# Diagnosing Influenza Infection from Pharyngeal Images using Deep Learning: Machine Learning Approach

**DOI:** 10.1101/2022.07.19.22276126

**Authors:** Sho Okiyama, Memori Fukuda, Masashi Sode, Wataru Takahashi, Masahiro Ikeda, Hiroaki Kato, Yusuke Tsugawa, Masao Iwagami

## Abstract

**Background:** Influenza is a major global burden of disease, causing annual epidemics and occasionally, pandemics. Given that influenza primarily infects the upper respiratory system, influenza infection may be able to be diagnosed by applying deep learning to pharyngeal images.

**Objective:** We aimed to develop a deep learning model to diagnose influenza infection using the data on pharyngeal images and clinical information.

**Methods:** We recruited patients who visited clinics and hospitals due to influenza-like symptoms. In the training stage, we developed a diagnostic prediction artificial intelligence (AI) model based on deep learning to predict polymerase chain reaction (PCR)-confirmed influenza from pharyngeal images and clinical information. In the validation stage, we assessed the diagnostic performance of the AI model. In the additional analysis, we compared the diagnostic performance of the AI model with that of three physicians, and also interpreted the AI model using the importance heatmaps.

**Results:** A total of 7,831 patients were enrolled at 64 hospitals between Nov 1, 2019 and Jan 21, 2020 in the training stage, and 659 patients (including 196 patients with PCR-confirmed influenza) at 11 hospitals between Jan 25, 2020 and Mar 13, 2020 in the validation stage. The area under the receiver operating characteristic curve of the AI model was 0.90 (95% confidence interval, 0.87–0.93), and its sensitivity and specificity were 76% (70–82%) and 88% (85–91%), respectively, outperforming three physicians. In the importance heatmaps, the AI model often focused on follicles on the posterior pharyngeal wall.

**Conclusions:** We developed the first AI model that can accurately diagnose influenza from pharyngeal images, which has the potential to assist physicians make timely diagnosis.

## Introduction

According to the Global Burden of Disease Study 2016, influenza is a major global burden of disease estimated to be the cause of 39.1 million acute lower respiratory infection episodes and 58,200 deaths [1]. It is estimated that influenza is responsible for 291,243 to 645,832 seasonal respiratory deaths (4.0–8.8 per 100,000 individuals) occur annually [2]. The timely and accurate diagnosis of influenza has the potential to prevent wide transmission of virus within the population and subsequent epidemic and pandemic, as well as the unnecessary prescription of antibiotics in primary care, which is a cause of emerging antibiotic-resistant bacteria. Moreover, early interventions such as hydration and antiviral drugs are expected to reduce the mortality risk among high-risk patients, including the elderly and individuals with comorbidities.

Although there are various tests currently used to diagnose influenza infection, the COVID-19 pandemic and the surge in the use of telemedicine highlighted the importance of accurately diagnosing it without increasing the risk of spreading virus through physical interactions. The gold-standard method of the diagnosis of influenza infection is the reverse-transcription polymerase chain reaction (RT-PCR) of nasopharyngeal aspirates or swabs [3, 4]; however, RT-PCR is not easily performed in primary care, and time to results could hamper the timely diagnosis and preventive/treatment interventions. A more commonly used test is the rapid immunochromatographic antigen detection tests, but compared with RT-PCR, their validity is modest and varies across studies [5, 6]. Both of these tests cannot be performed through telemedicine, whereas the sensitivity and specificity of diagnosing influenza only with clinical information are suboptimal [7, 8]. Given that more patients are diagnosed through telemedicine in recent years, an alternative test of influenza that could be conducted through telemedicine is warranted.

To address this important knowledge gap, we developed a deep learning model to diagnose influenza infection using the data on pharyngeal images and clinical information. We tested the performance of the diagnostic prediction artificial intelligence (AI) model using the data on the real-world patient population, and also compared it with the diagnostic performance of three physicians. We also investigated the regions of the pharynx on which the AI model focused to differentiate between individuals with and without influenza infection.

## Methods

### Pilot study to develop a medical camera to capture standardized pharyngeal images

For our pilot study, we recruited 4,765 patients aged 6–90 years with influenza-like symptoms who visited 37 clinics or hospitals between Nov 28, 2018 and Feb 4, 2019 (registered as jRCTs032180041). We developed a pharyngeal camera with a light emitting diode light source and a disposable clear cap to hold down the tongue of patients, to capture images of the pharynx in a standardized manner (**Figure 1**). In this pilot study, we adjusted the size of the pharyngeal camera and tongue depressors to be suitable for many patients. The device contains a full high-definition digital camera and is connected via Wi-Fi to a cloud service for the analysis of the pharyngeal images, together with clinical information. We also improved the image quality of the camera, such as resolution, brightness, and contrast, during this pilot study. We used a rapid continuous shooting function to obtain high-quality pharyngeal images in a short time by avoiding motion blur. The camera can capture an image every 0.3 seconds, and 30 sequential images are captured per shot.

**Figure 1.**
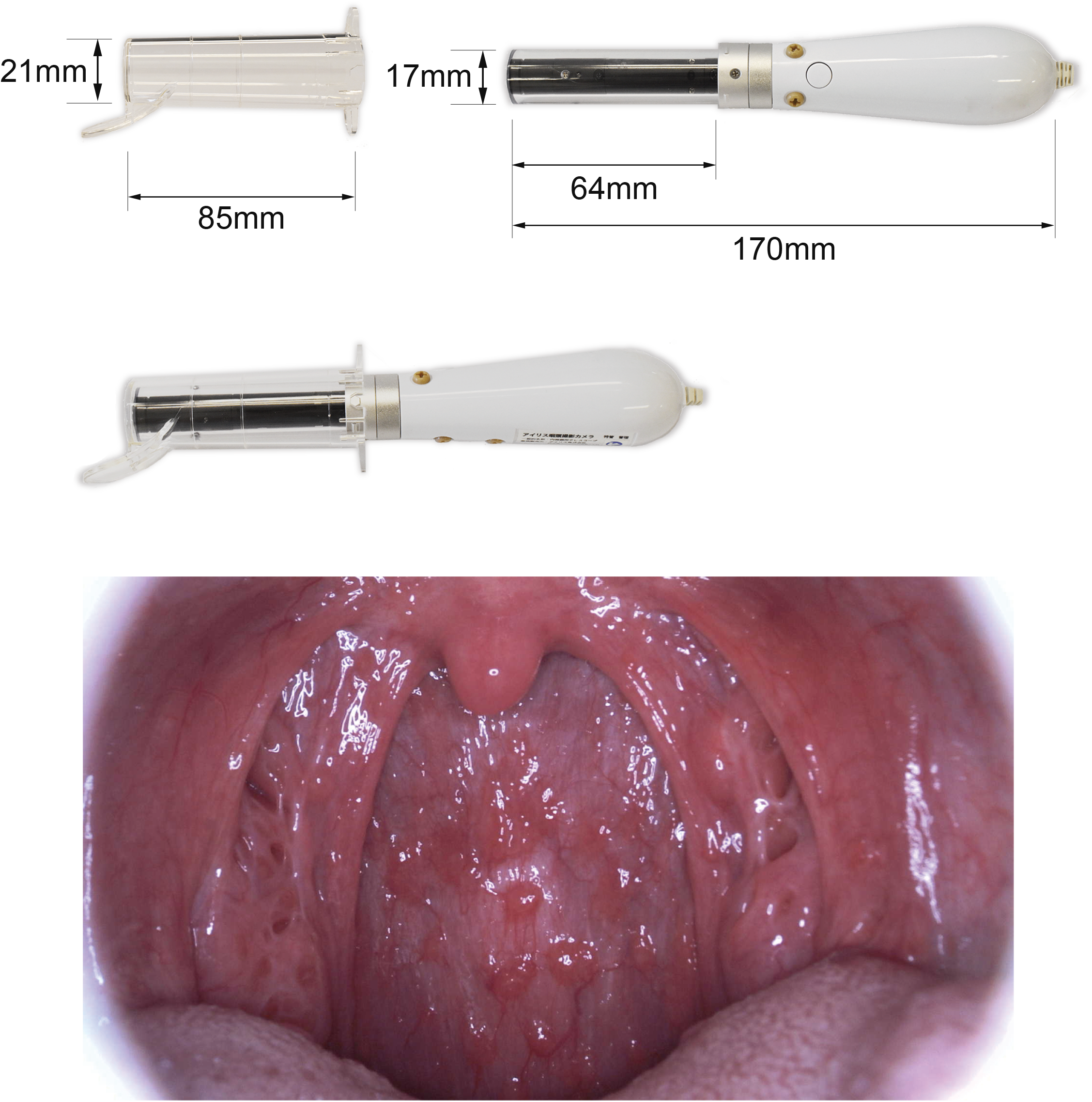
Presentation of the AI-assisted camera and a representative pharyngeal image of a patient with PCR-confirmed influenza infection. AI: artificial intelligence, PCR: polymerase chain reaction.

### Study design and participants

The current study included a training stage (registered as jRCTs032190120) and validation stage (registered as Pharmaceuticals and Medical Devices Agency clinical trial identification code AI-02-01). We enrolled patients with influenza-like symptoms who visited clinics or hospitals and satisfied the following inclusion and exclusion criteria at 64 hospitals between Nov 1, 2019 and Jan 21, 2020 in the training stage, and 11 hospitals between Jan 25, 2020 and Mar 13, 2020 in the validation stage. The list of study sites is shown in the **Supplementary Table 1**.

The inclusion criteria were (i) patients with written consent to participate in the study provided by themselves or their parent (if they were younger than 18 years), (ii) those aged six years or older, and (iii) those that satisfied at least one of the following four conditions in the training stage and at least two in the validation stage: (a) body temperature of 37.0°C or higher, (b) systematic influenza-like symptoms, such as joint pain, muscle pain, headache, tiredness, and appetite loss, (c) respiratory symptoms, such as cough, sore throat, and nasal discharge or congestion, and (d) an episode of close contact with patients with influenza or influenza-like symptoms within three days, or in any other scenario in which the consulting physician suspected influenza infection. The exclusion criteria included (i) those with fluctuating teeth, (ii) those with severe oral lesions, (iii) those with severe nausea, (iv) those with difficulty in opening the mouth sufficiently for the use of the camera (e.g., small mouth, temporomandibular joint pain, incompatibility of dentures, disturbed consciousness, or respiratory failure), (v) those who had participated in another clinical trial within seven days prior, those who were scheduled to participate in another clinical trial (excluding post-marketing surveillance), or those with difficulty in follow-up for mental, family, social, geographical, or other reasons, (vi) pediatric patients who clearly did not agree to participate in the study, and (vii) those judged to be inappropriate to participate in the study by the responsible physician at each site. Additionally, patients with only poor-quality images were excluded from the analysis.

In the training stage, we aimed to collect clinical information and pharyngeal images of PCR-confirmed influenza positive and negative patients in a ratio of roughly 1:1 for the most efficient supervised learning of the AI model. There is no consensus on the size of samples (i.e., number of patients) that should be used to train an AI model; thus, we arbitrarily set them to 7,500 patients, including 3,750 influenza PCR-positive and 3,750 PCR-negative. In the validation stage, we aimed to determine the lower bound of the 95% one-sided confidence interval (CI) of sensitivity to achieve 70% or higher, and that of specificity to achieve 85% or higher. With a one-sided p value of 5% and power of 85%, assuming an actual sensitivity of 80% and specificity of 90% as suggested by our training stage, we calculated the required sample sizes to be 137 for influenza PCR-positive cases and 323 for PCR-negative cases. Therefore, we planned to stop the recruitment of the study participants on the day that both 150 positive cases and 350 negative cases were obtained.

In Japan, the first case of the SARS-CoV-2 infection (COVID-19) was reported on Jan 15, 2020, and the first wave of the pandemic occurred from late March 2020. During the study period, in the validation stage, we asked the participating clinics or hospitals to report any suspected cases of COVID-19 in the study participants. There were no such reports from any study site throughout the research, which suggests that our study was not affected by the COVID-19 pandemic.

### Collection of pharyngeal images, clinical information, and nasopharyngeal specimens

In addition to the pharyngeal images of the study participants, the following clinical information was obtained using a standardized case report form based on electronic data capture: age; sex; time (hours) from symptom onset; highest body temperature before study site visit; episode of close contact; status and date of the most recent influenza vaccination; use of antipyretics; subjective symptoms, including tiredness, appetite loss, chill, sweating, joint pain, muscle pain, headache, nasal discharge or congestion, cough, sore throat, and digestive symptoms; and objective findings at the study sites, including body temperature, pulse rate, and tonsillar findings (tonsillitis, white moss, and redness) according to the consulting physician.

Furthermore, nasopharyngeal swabbing was conducted to obtain nasopharyngeal specimens from the participants, which were sent to the central clinical laboratory (LSI Medience Corporation, Tokyo, Japan) for RT-PCR, which is the gold standard (reference standard) for diagnosis of influenza infection. We standardized the process of collecting the nasopharyngeal specimens among the study sites using a manual.

### Development of the AI model to predict PCR-confirmed influenza

An ensemble AI model (version 2.0) was developed to predict the probability of PCR-confirmed influenza using pharyngeal images and clinical information. This model consisted of three main machine learning models: a multi-view convolutional neural network (MV-CNN), multi-modal convolutional neural network (MM-CNN), and boosting models. The three models were trained on the same training dataset from the training stage and combined using ridge regression [9].

First, the MV-CNN was trained using SE-ResNext-50 as an image feature extractor pre-trained on ImageNet [10, 11]. The MV-CNN architecture used several pharyngeal images that contained views from various angles [12]. In pharyngeal imaging, the tongue and uvula often overlap with the posterior pharyngeal wall. The MV-CNN addressed this issue by gathering information from various image angles. One to five of the most appropriate images per patient were selected by an automatic image quality evaluation system using a lightweight CNN model [13]. The system was trained using image quality criteria defined by a physician among the authors (MF) in the training stage. The input images for the MV-CNN were resized and then augmented (e.g., flipped, rotated, blurred, and contrast changed) to improve accuracy and generalization performance. To prevent overfitting, well-established training strategies were used, including batch normalization, learning rate decay, and cross-validation. To manage various pharyngeal magnification rates, MV-CNNs with multiple image sizes were trained and their scores were combined by averaging them.

Second, the MM-CNN was developed based on the MV-CNN to process both multi-view pharyngeal images and clinical information as input data [14, 15]. In detail, the final classification layer of the MV-CNN was extended and connected to the neural network to manage clinical information. The image feature extractor of the MM-CNN was initialized with the trained MV-CNN weights. Then, the same training and ensemble strategy as the MV-CNN were applied.

Third, boosting models were trained based on the prediction results of the MV-CNN and clinical information. LightGBM and CatBoost were selected as boosting models [16, 17]. Finally, the probability of influenza was obtained by integrating each prediction from the MV-CNN, MM-CNN, and boosting models using ridge regression. The ridge regression parameters were determined using cross-validation.

### Statistical analysis

In the training stage, we compared the clinical characteristics of the study participants according to the PCR test result (positive or negative) using t-tests for continuous variables with a normal distribution (age, highest body temperature before the study site visit, body temperature at visit, and pulse rate), the Mann–Whitney U test for continuous variables with a non-normal distribution (time from symptom onset), and chi-square tests for categorical variables. We repeated these analyses in the validation stage.

In the training stage, using a five-fold cross-validation method, we conducted receiver operating characteristic (ROC) curve analysis to measure the discrimination ability of (i) the probability score of the MV-CNN, which uses only pharyngeal images in prediction, (ii) the probability score of the clinical information AI, which is an AI model that uses all the other aforementioned clinical information (except for the pharyngeal images) in prediction, and (iii) the probability score of the ensemble AI model using both the pharyngeal images and clinical information. We also measured the reclassification ability of the pharyngeal images by comparing the clinical information AI model and the ensemble AI model by calculating the continuous net reclassification improvement (NRI) and integrated discrimination improvement (IDI) [18].

In the validation stage, we also conducted ROC analysis and calculated the sensitivity, specificity, positive predictive value (PPV), and negative predictive value (NPV) for influenza infection, according to a selected cut-off point.

We performed statistical analysis using R (version 4.1.1) and Python (version 3.8.5). P values of < 0.05 were considered to be statistically significant. A third-party organization (Statcom Co., Ltd., Tokyo, Japan) performed the sample size estimation, and calculation of the area under the receiver operating characteristic curve (AUROC) and validity (sensitivity, specificity, PPV, and NPV) in the validation stage.

### Additional analysis

We conducted two types of additional analysis. First, we compared the performance of the AI-assisted diagnosis camera with that of three physicians. For this analysis, we used the existing data (pharyngeal images and clinical information) of 200 patients (100 influenza PCR-positive cases and 100 PCR-negative cases), which was randomly selected from the study participants in the training stage. Three physicians among the authors (SO, MF, and MIk), who were blinded to patients’ identifiers and their PCR test results, assessed the data to give an influenza prediction score between 0 and 1 (i.e., between 0% and 100%). We applied the diagnostic prediction AI model to these existing data, and compared the AUROC of the diagnostic prediction AI model with that of each physician and the average prediction score of the three physicians. We recalculated the AUROC of the AI model for these 200 patients for a fair comparison.

Second, we attempted to interpret the mechanisms of the MV-CNN prediction to differentiate between influenza cases and non-cases from the pharyngeal images. We modified guided gradient-weighted class activation mapping (Guided Grad-CAM) for the MV-CNN to visualize importance heatmaps. The aim was to show where the MV-CNN focused when differentiating between influenza PCR-positive cases and PCR-negative cases. We used the same dataset of 200 patients (100 PCR-positive cases and 100 PCR-negative cases) as the first additional analysis. To quantify and interpret the importance heatmaps, two physicians among the authors (MF and MIk) independently determined whether the MV-CNN highlighted each part of the pharynx (classified into five parts: lateral pharyngeal bands, posterior pharyngeal wall, palatal arch, tonsils, and follicles) for each patient. When the two physicians made different judgements (i.e., presence vs. absence of highlighting by the MV-CNN), consensus was reached through discussion between them. Consequently, for each part of the pharynx, we calculated the proportion of patients with images highlighted by the MV-CNN among the 100 PCR-positive cases and 100 PCR-negative cases, and compared the groups using chi-square tests.

## Results

### The training stage

In the training stage, we obtained informed consent from 9,029 patients with influenza-like symptoms who visited one of 64 clinics or hospitals between Nov 1, 2019 and Jan 21, 2020. Among them, 199 patients (2.2%) felt nauseous during the examination when the pharyngeal images were being captured, including one patient with severe nausea and 14 patients (0.16%) who vomited. We did not complete the image-capturing procedure for these 15 patients (0.16%). Among the remaining 9,014 patients, we selected 7,831 patients (mean age 33.8 years [SD 18.4 years], women 50%) with 25,168 high-quality images (out of approximately 300,000 images), which consisted of 3,733 influenza PCR-positive patients with 12,154 pharyngeal images and 4,098 PCR-negative patients with 13,014 pharyngeal images. Table 1 compares the clinical characteristics of patients by PCR test results. Compared with the PCR-negative cases, the PCR-positive cases yielded the following results: the average age was slightly lower; time from symptom onset to the study site visit was shorter; the proportion of close contact, use of antipyretics, and most subjective symptoms were higher; and the temperature and pulse rate were higher, whereas the proportion of recent influenza vaccinations, digestive symptoms, and tonsillar findings were lower. There was no difference in the proportion of sex and sore throat between the groups.

**Table 1.** Characteristics of study participants with or without RT-PCR confirmed influenza.

|  | Participants in the training stage |  |  |  | Participants in the validation stage |  |  |  |
| --- | --- | --- | --- | --- | --- | --- | --- | --- |
|  | All<br>(n=7831) | by RT-PCR test result |  |  | All<br>(n=659) | by RT-PCR test result |  |  |
|  |  | Positive<br>(n=3733) | Negative<br>(n=4098) | p<br>value |  | Positive<br>(n=196) | Negative<br>(n=463) | p<br>value |
| Age (years), mean±SD | 33.8±18.4 | 33.0±18.5 | 34.5±18.4 | <0.001 | 33.3±17.6 | 30.4±18.6 | 34.5±17.0 | 0.009 |
| Sex: |  |  |  | 0.539 |  |  |  | 0.542 |
| Male | 3930 (50.2%) | 1887 (50.5%) | 2043 (49.9%) |  | 318 (48.3%) | 91 (46.4%) | 227 (49.0%) |  |
| Female | 3901 (49.8%) | 1846 (49.5%) | 2055 (50.1%) |  | 341 (51.7%) | 105 (53.6%) | 236 (51.0%) |  |
| Time from onset (hours), mean±SD | 31.2±25.3 | 28.3±20.6 | 33.8±28.6 | <0.001 | 27.5±31.2 | 24.6±10.8 | 28.7±36.5 | 0.671 |
| Highest BT before visit (°C), mean±SD | 38.2±0.9 | 38.6±0.8 | 38.0±0.9 | <0.001 | 38.2±0.8 | 38.6±0.7 | 38.0±0.8 | <0.001 |
| Close contact | 2520 (32.2%) | 1687 (45.2%) | 833 (20.3%) | <0.001 | 208 (31.6%) | 120 (61.2%) | 88 (19.0%) | <0.001 |
| Recent flu vaccination | 2873 (36.7%) | 1248 (33.4%) | 1625 (39.7%) | <0.001 | 278 (42.2%) | 73 (37.2%) | 205 (44.3%) | 0.095 |
| Use of antipyretics | 2975 (38.0%) | 1530 (41.0%) | 1445 (35.3%) | <0.001 | 297 (45.1%) | 95 (48.5%) | 202 (43.6%) | 0.254 |
| Subjective symptoms: |  |  |  |  |  |  |  |  |
| Tiredness | 5937 (75.8%) | 3010 (80.6%) | 2927 (71.4%) | <0.001 | 506 (76.8%) | 159 (81.1%) | 347 (74.9%) | 0.086 |
| Appetite loss | 3361 (42.9%) | 1823 (48.8%) | 1538 (37.5%) | <0.001 | 259 (39.3%) | 96 (49.0%) | 163 (35.2%) | <0.001 |
| Chill | 4215 (53.8%) | 2231 (59.8%) | 1984 (48.4%) | <0.001 | 338 (51.3%) | 115 (58.7%) | 223 (48.2%) | 0.014 |
| Sweating | 2188 (27.9%) | 1128 (30.2%) | 1060 (25.9%) | <0.001 | 206 (31.3%) | 60 (30.6%) | 146 (31.5%) | 0.816 |
| Joint pain | 3735 (47.7%) | 1992 (53.4%) | 1743 (42.5%) | <0.001 | 316 (48.0%) | 103 (52.6%) | 213 (46.0%) | 0.124 |
| Muscle pain | 2362 (30.2%) | 1276 (34.2%) | 1086 (26.5%) | <0.001 | 192 (29.1%) | 62 (31.6%) | 130 (28.1%) | 0.359 |
| Headache | 4725 (60.3%) | 2414 (64.7%) | 2311 (56.4%) | <0.001 | 403 (61.2%) | 126 (64.3%) | 277 (59.8%) | 0.283 |
| Nasal discharge or congestion | 4472 (57.1%) | 2202 (59.0%) | 2270 (55.4%) | 0.001 | 410 (62.2%) | 134 (68.4%) | 276 (59.6%) | 0.034 |
| Cough | 5219 (66.6%) | 3053 (81.8%) | 2166 (52.9%) | <0.001 | 384 (58.3%) | 161 (82.1%) | 223 (48.2%) | <0.001 |
| Sore throat | 4928 (62.9%) | 2353 (63.0%) | 2575 (62.8%) | 0.857 | 440 (66.8%) | 126 (64.3%) | 314 (67.8%) | 0.379 |
| Digestive symptoms | 1298 (16.6%) | 558 (14.9%) | 740 (18.1%) | <0.001 | 127 (19.3%) | 30 (15.3%) | 97 (21.0%) | 0.093 |
| Objective findings: |  |  |  |  |  |  |  |  |
| BT at visit (°C) mean±SD | 37.6±0.9 | 38.0±0.9 | 37.3±0.8 | <0.001 | 37.5±0.9 | 37.9±0.9 | 37.3±0.8 | <0.001 |
| Pulse rate, mean±SD | 95.0±17.8 | 100.2±17.7 | 90.3±16.6 | <0.001 | 93.8±17.7 | 100.8±18.6 | 90.9±16.4 | <0.001 |
| Tonsillitis | 1238 (15.8%) | 529 (14.2%) | 709 (17.3%) | <0.001 | 63 (9.6%) | 8 (4.1%) | 55 (11.9%) | 0.002 |
| Tonsillar white moss | 126 (1.6%) | 17 (0.5%) | 109 (2.7%) | <0.001 | 23 (3.5%) | 1 (0.5%) | 22 (4.8%) | 0.007 |
| Tonsillar redness | 1292 (16.5%) | 540 (14.5%) | 752 (18.4%) | <0.001 | 69 (10.5%) | 13 (6.6%) | 56 (12.1%) | 0.036 |

Abbreviations: BT: body temperature, SD: standard deviation, RT-PCR: real-time reverse transcription polymerase chain reaction

Using the training dataset, we established the ensemble AI model to estimate the probability of influenza for individual patients. The feature importance of each variable in the LightGBM model and CatBoost model is shown in the **Supplementary Figures 1 and 2**, which suggest that pharyngeal images were the most important variable in the diagnostic prediction AI model, followed by body temperature and cough.

In the five-fold cross-validation, the AUROC of the MV-CNN probability score for pharyngeal images was 0.76 (95% CI 0.75–0.77) and that of the AI model with clinical information (i.e., all the clinical information in **Table 1**) was 0.83 (95% CI 0.82–0.84) (**Figure 2**). Taken together, the AUROC of the diagnostic prediction AI model was 0.87 (95% CI 0.86–0.87), which means that the AUROC significantly increased as a result of adding pharyngeal images to the AI model with clinical information (p < 0.001). Regarding reclassification ability, the continuous NRI was 0.25 (95% CI 0.22–0.29) among PCR-positive cases and 0.33 (95% CI 0.30–0.36) among PCR-negative cases, and IDI was 0.08 (95% CI 0.07–0.08), which also indicates that the accuracy of the diagnostic prediction AI model significantly improved as a result of adding pharyngeal images to the AI model with clinical information.

**Figure 2.**
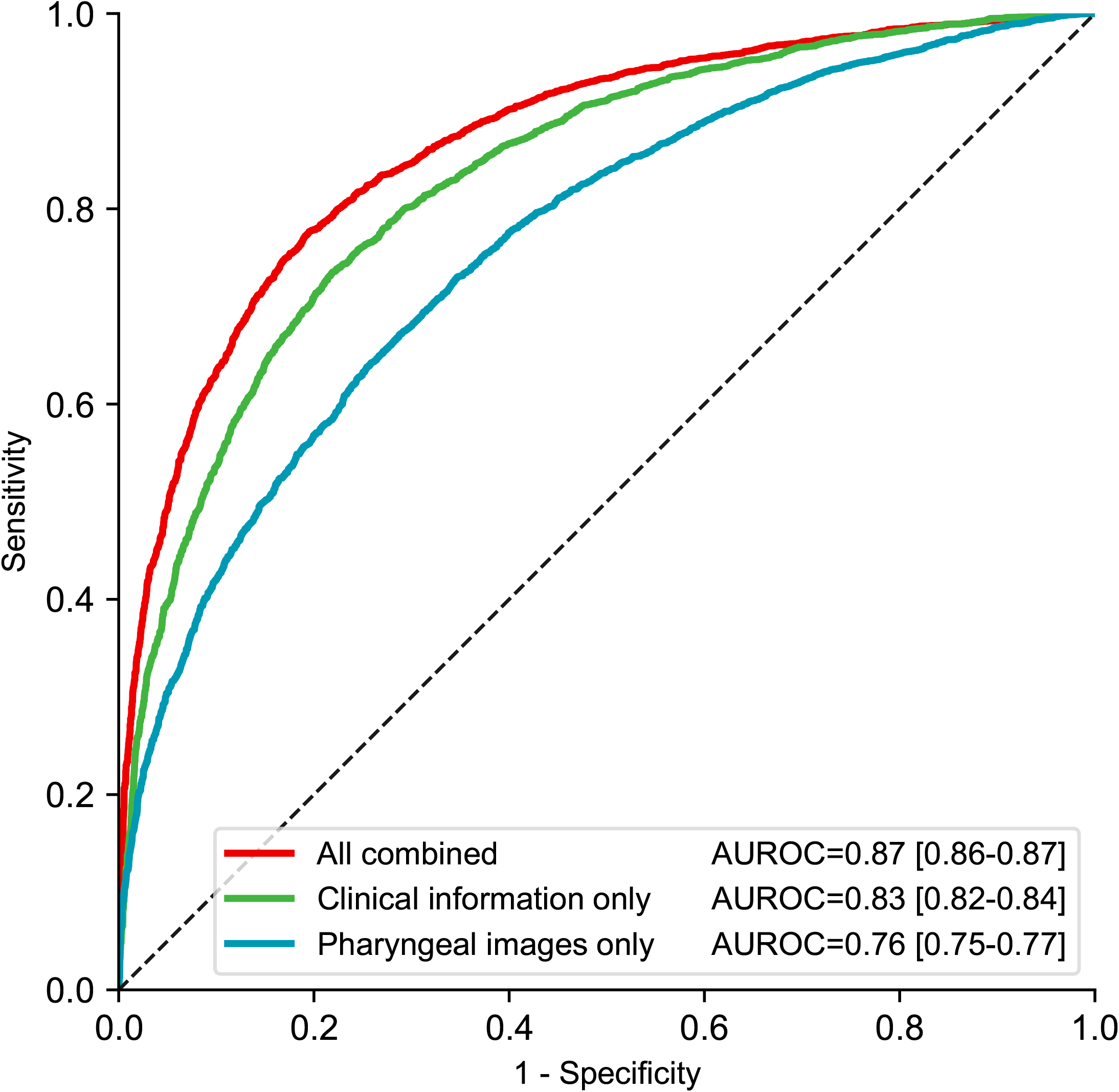
Receiver operating characteristic curves of the diagnostic prediction models in the five-fold cross-validation of the training dataset. AUROC: area under the receiver operating characteristic curve, All combined: ensemble AI model using pharyngeal images and clinical information, Pharyngeal images only: multi-view convolutional neural network using multiple pharyngeal images, Clinical information only: ensemble AI model without pharyngeal image information.

### The validation stage

In the validation stage, we obtained informed consent from 706 patients with influenza-like symptoms who visited one of 11 clinics or hospitals between Jan 25, 2020 and Mar 13, 2020, which comprised a safety analysis set. Among them, 12 patients (1.7%) felt nauseous during the examination when the pharyngeal images were being captured, including one patient (0.1%) with severe nausea for whom we did not complete the image-taking procedure. Additionally, 33 patients (4.7%) did not satisfy the predefined criteria of the protocol for the full analysis set, mostly because of trouble in saving the pharyngeal images at the study sites. Furthermore, 13 patients were excluded by the automated image quality evaluation system that removed low-quality pharyngeal images. Thus, we used the pharyngeal images and clinical information of the remaining 659 patients (mean age 33.3 years [SD 17.6 years], women 51.7%) for the following analysis. Similar to the training stage, compared with non-cases, the PCR-confirmed cases yielded the following results: the average age was slightly lower, the proportion of close contact and several subjective symptoms (tiredness, chill, nasal discharge/obstruction, and cough) was higher, and the temperature (both before the clinic/hospital visit and on site) and pulse rate were higher, whereas the proportion of tonsillar findings was lower (**Table 1**).

In the validation stage, the AUROC of the diagnostic prediction AI model was 0.90 (95% CI 0.87-0.93). At a selected cut-off point on the ROC (**Supplementary Figure 3**), the sensitivity and specificity were 76% (95% CI 70-82%) and 88% (95% CI 85-91%), and the PPV and NPV were 73% (95% CI 69-79%) and 90% (95% CI 87-92%), respectively (**Supplementary Table 2**).

### The additional analysis

In our additional analysis, among the 200 randomly selected patients (100 influenza PCR-positive cases and 100 PCR-negative cases), the AUROC of the diagnostic prediction AI model was 0.89 (95% CI 0.84–0.93) and was higher than that of each of the three physicians (0.76, 0.73, and 0.74, respectively). It was also higher than that of the average prediction score of the three physicians (0.79, 95% CI 0.73–0.85) (**Figure 3**).

**Figure 3.**
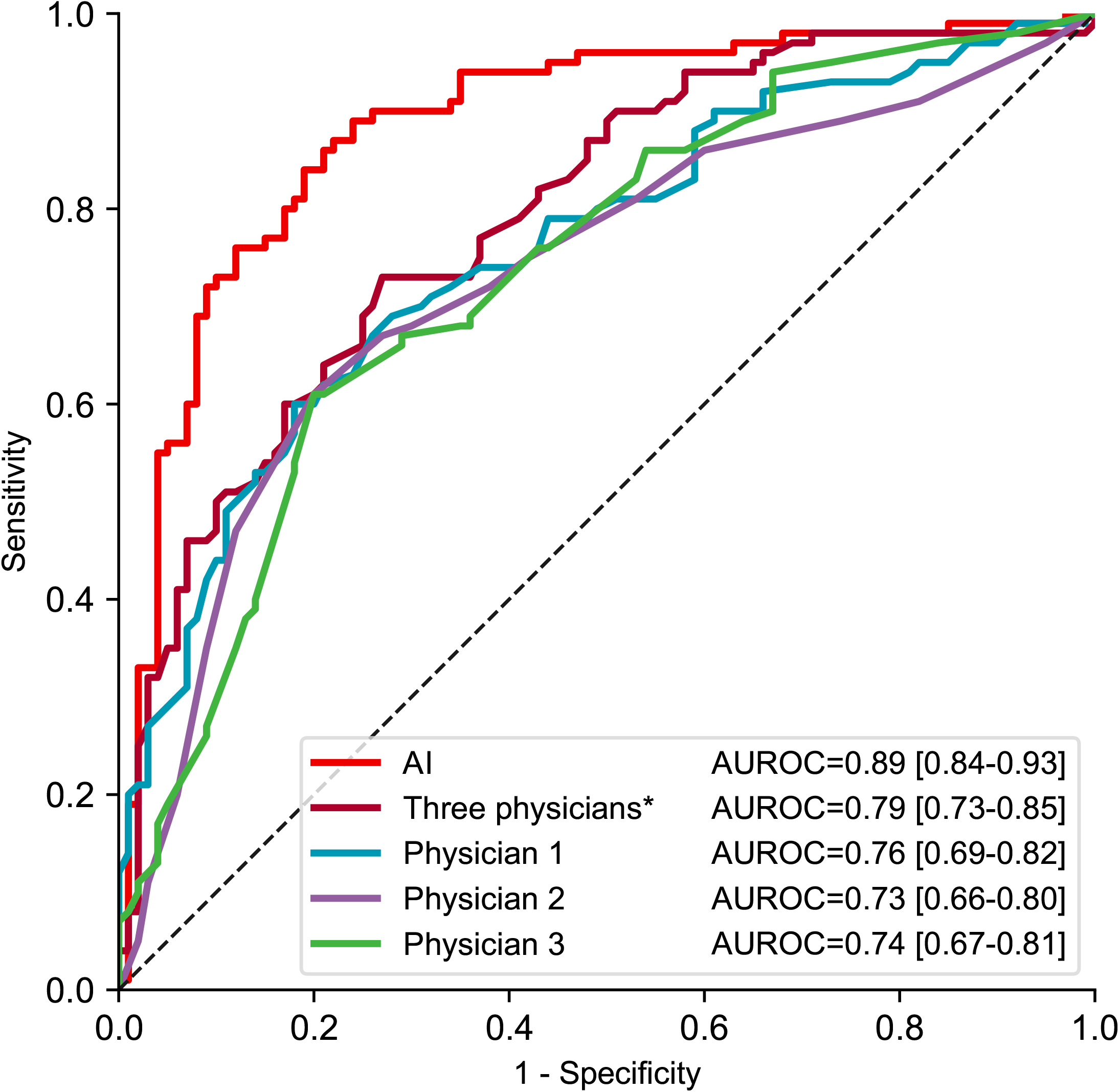
Receiver operating characteristic curves for the diagnostic prediction AI model and three physicians. AUROC: area under the receiver operating characteristic curve, AI: ensemble AI model using pharyngeal images and clinical information, *average prediction score of three physicians Note: The AI model is the same as that used in the validation stage. However, the AUROC is slightly different because of the small sample size used in the additional analysis.

**Figure 4** shows examples of the pharyngeal images and those highlighted using the importance heatmaps for three patients. An assessment of the importance heatmaps for the 200 patients (100 PCR-positive cases and 100 PCR-negative cases) conducted by two physicians showed that the proportion of patients with images highlighted by the AI model was significantly different between the PCR-positive cases and PCR-negative cases for follicles on the posterior pharyngeal wall (73% vs. 38%, p < 0.001), which suggests that the AI model often focused on these parts (**Figure 5**).

**Figure 4.**
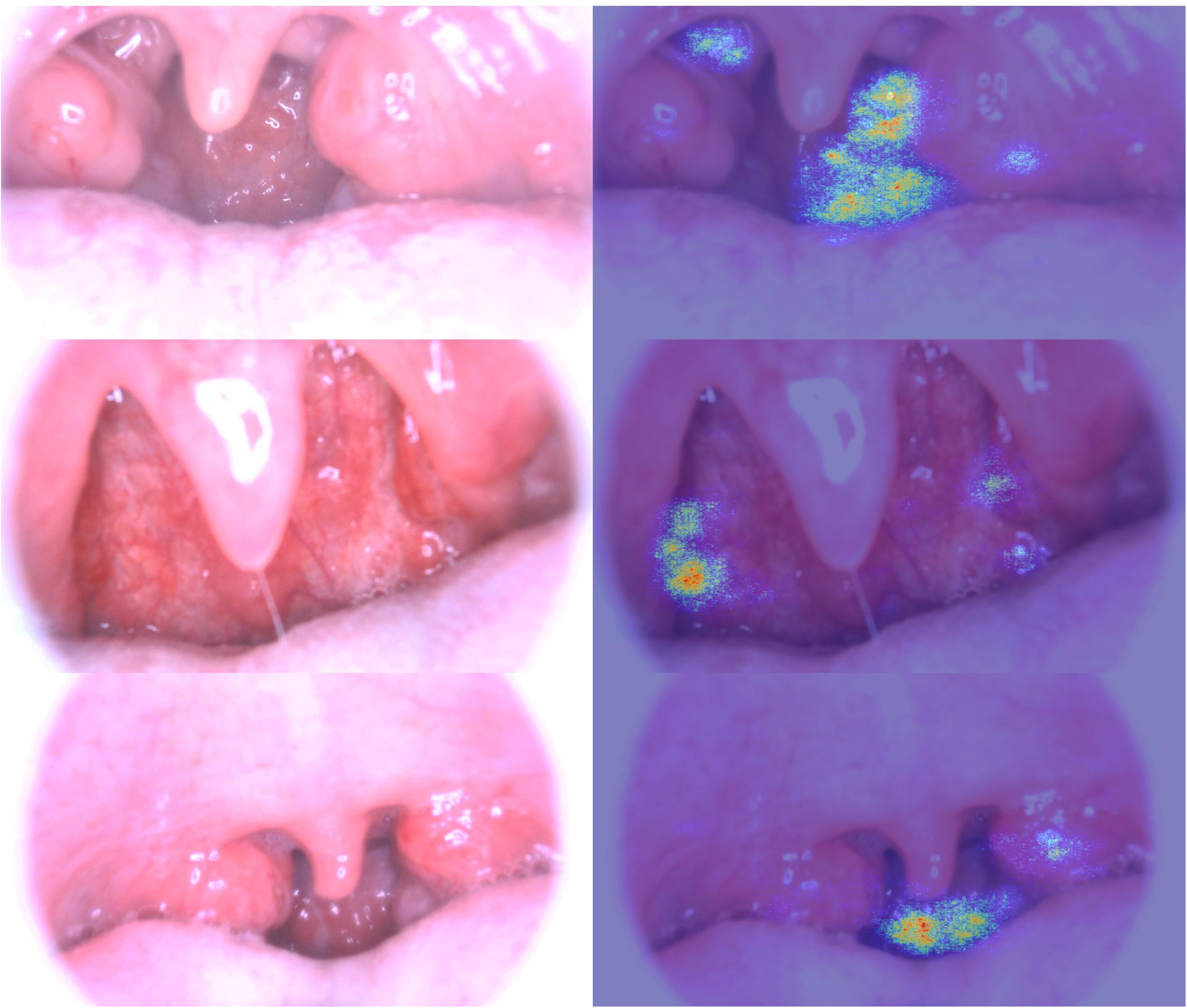
Examples of pharyngeal images (left) and those highlighted using the importance heatmaps (right) for three patients. Note: These importance heatmaps show areas that the AI model focused on to differentiate between the PCR-positive cases and PCR-negative cases.

**Figure 5.**
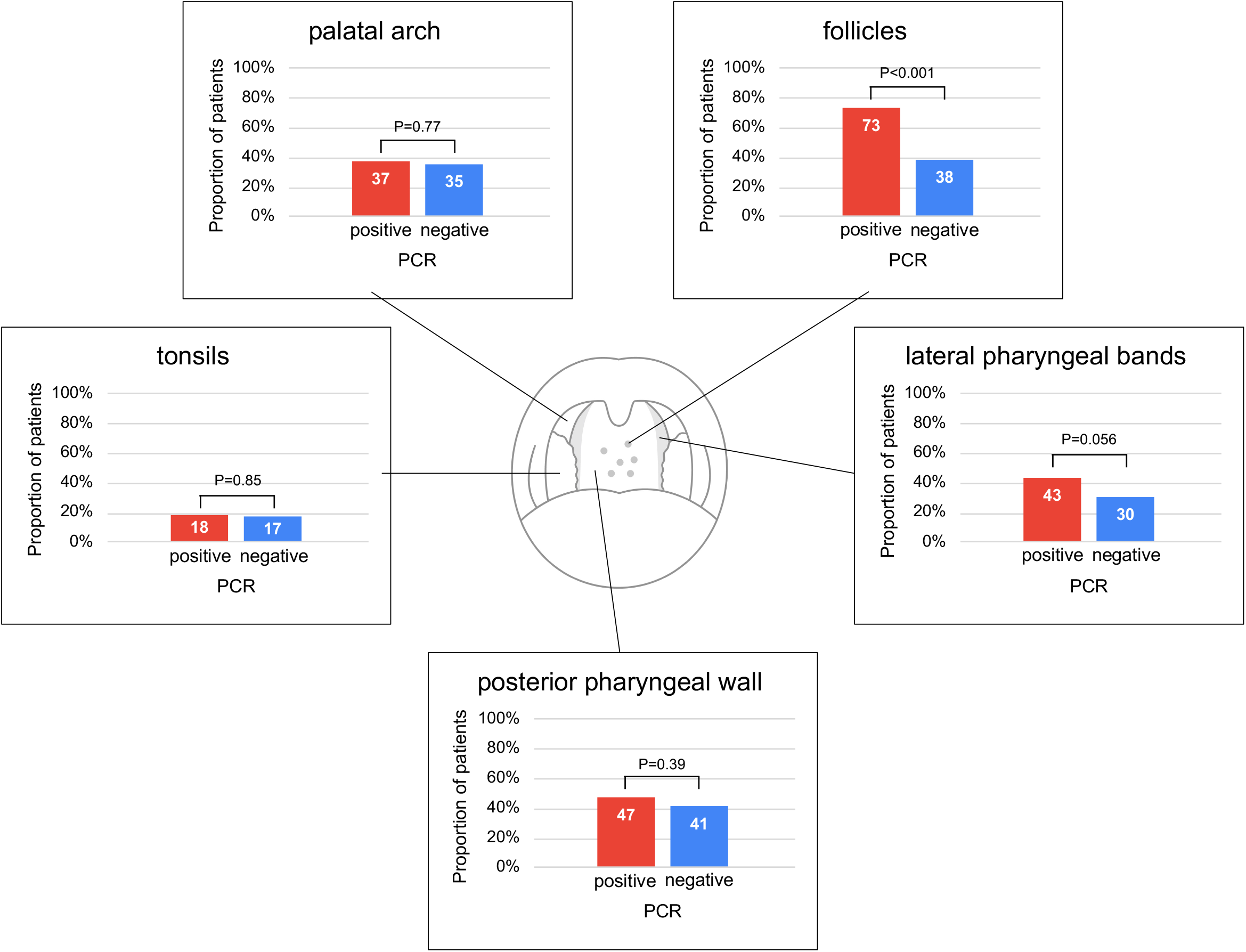
Proportion of patients with images highlighted by the AI model on each part of the pharynx in 100 PCR-positive influenza cases and 100 PCR-negative cases. PCR: polymerase chain reaction.

## Discussion

In this study, we developed an AI-assisted diagnosis camera with a diagnostic prediction model for influenza. In the training stage, we found that the pharyngeal images contributed significantly to the improvement of the diagnosis prediction AI model compared with the clinical information AI. In the validation stage, the AUROC of the diagnostic prediction AI model was 0.90 (95% CI 0.87-0.93), with a sensitivity and specificity of 76% (95% CI 70-82%) and 88% (95% CI 85-91%), respectively. In our additional analysis, the AI-assisted camera performed better than three physicians in predicting influenza. Furthermore, in the importance heatmaps, we found that the AI model often focused on follicles to differentiate between PCR-positive and negative cases.

Clinical characteristics associated with PCR-confirmed influenza infection among people with influenza-like symptoms were examined in two previous studies [7, 8]. Both of these studies concluded that fever and cough were the best predictors of influenza diagnosis. However, the sensitivity and specificity of the combination of these two factors were suboptimal, at 78% and 55% in one study [7], and 64% and 67% in another study, respectively [8]. In our study, considering the feature importance of each variable in the LightGBM model and CatBoost model (**Supplementary Figures 1 and 2**), body temperature and cough were highly ranked among the clinical information, whereas the feature importance of pharyngeal images was even larger.

Recently, several AI-assisted diagnostic prediction models have been proposed for influenza diagnosis [19-22]. A single-center study from Japan reported a machine learning-based infection screening system incorporating a random tree algorithm that used vital signs [19]. The researchers reported a sensitivity of 81–96% and NPV of 81–96% in their training datasets (specificity and PPV were not reported), but the performance of the model was not validated outside the center. The University of Pittsburgh Medical Center Health System reported machine learning classifiers for influenza detection from emergency department free-text reports [20-21]. Among the 31,268 emergency department reports from four hospitals, the AUROCs of the seven machine learning classifiers for influenza detection ranged from 0.88 to 0.93 [21], which was better than an expert-built Bayesian model [20]. These studies were also limited because performance outside the health care system of the University of Pittsburgh was unknown. More recently, a Korean study reported an influenza screening system based on deep learning using a combination of epidemiological and patient-generated health data from a mobile health app [22]. However, the gold standard in the study was the clinical diagnosis of influenza at a clinic reported by app users instead of laboratory-based confirmed influenza. Notably, none of the previous studies included an assessment of pharyngeal images in their diagnostic prediction models [19-22]. The novelty of our study is that we have developed the first AI-assisted diagnosis camera for influenza and prospectively validated its performance through a Good Clinical Practice-based clinical trial process.

We showed that pharyngeal images significantly improved the discrimination and reclassification ability of the diagnostic prediction AI model. Additionally, we considered the mechanisms by which the AI model differentiated between true influenza cases and non-influenza cases using pharyngeal images. To the best of our knowledge, there has been no established approach to quantitatively scale regions of images on which the AI focuses. Indeed, most previous studies on AI-assisted diagnosis cameras showed only representative images highlighted using Grad-CAM or saliency maps to speculate on possible mechanisms of AI classification [23-25]. In our study, we attempted to quantify these regions by calculating the proportion of patients with images highlighted by the AI model for each part of the pharynx among the influenza PCR-positive cases and negative cases. Consequently, we found that the AI model mainly focused on follicles on the posterior pharyngeal wall.

Notably, this finding is in line with previous case reports and case series that suggest that follicles on the posterior pharyngeal wall are specific to influenza infection and useful for the diagnosis of influenza [26-29]. Physical examination, including visual inspection of the pharynx, generally requires the experience of individual physicians, and physical examination skills may vary widely among physicians. Our study suggests that AI could minimize the variation and may help to standardize physical examination skills among physicians. Additionally, when attempting to discriminate between diseases, doctors may be able to learn from AI systems where to focus in their visual examination.

Our study has limitations. First, we recruited study participants with influenza-like symptoms from a large number of clinics and hospitals in Japan to increase the generalizability of our study. However, there may be a country or cultural difference in terms of people with influenza-like symptoms seeking medical care from healthcare providers. In Japan, with its universal health care coverage, people have relatively easy and timely access to clinics or hospitals compared with those in other countries. Therefore, generalizing our findings to different clinical care settings in different countries would require caution and may require independent assessment. Second, our additional analysis of the comparison between the AI-assisted diagnosis camera and three physicians was not pre-planned in the study protocols (jRCTs032190120 and Pharmaceuticals and Medical Devices Agency clinical trial identification code AI-02-01), although these physicians were blinded to patients’ identifiers and their PCR results. Finally, in addition to the pharyngeal images, we collected as many relevant clinical variables (suggested by previous large studies [7, 8]) as possible to establish an accurate diagnostic prediction AI model. However, there may be other useful variables for the prediction of true influenza diagnosis that were not collected in our study. For example, some studies have suggested that the population level trend of influenza outbreaks in an area is useful for predicting an individual patient’s influenza infection [22]. Further improvement of the AI-assisted diagnosis camera by including additional variables, in addition to an improvement of the AI models to analyze pharyngeal images, are justified.

In conclusion, we developed the first AI-assisted diagnosis camera for influenza and prospectively validated its high performance. We found that the AI model often focused on follicles, which confirmed previous case reports and series that suggested that visual inspection of the pharynx would help to diagnose influenza infection.

## Data Availability

The data used in this study, mainly pharyngeal images, are licensed to Aillis, Inc.. The data have not been opened in public, and could be used for future projects for the development of medical devices and diagnostic technologies. Proposals and requests for data access should be directed to the corresponding author via email.

## Acknowledgements

We thank all the study participants, medical staff at the research facilities, and physicians, particularly Dr. Jun Fukuda. We would also like to thank Shuhei Fujimoto and Tetsuro Oda for their statistical advice, and Dr. Akihiko Miyamoto and Dr. Shigeyuki Watanabe for their clinical advice. We trained the AI model on the AI Bridging Cloud Infrastructure of the National Institute of Advanced Industrial Science and Technology (AIST), Japan. We thank A2 Healthcare Corporation, Tokyo, Japan, for assisting with the clinical study operation, data collection process, auditing, and quality control and assurance, and Statcom Co., Ltd., Tokyo, Japan, for performing statistical analysis in the validation stage. The development of the camera was led by members of the hardware team of Aillis, including Takashi Yasumi; the development of the AI model was led by members of the AI team, including Atsushi Fukuda; and the clinical studies were led by the medical team, including Miho Nakamura. All the members of Aillis and their dedicated efforts have made this study possible, including, but not limited to, the aforementioned teams.

## Funding

This study was funded by the New Energy and Industrial Technology Development Organization (NEDO), Japan (grant number 30STS713) and Aillis, Inc. The NEDO had no role in the study design, data collection, data analysis, data interpretation, or writing of the report. The role of Aillis, Inc. in the pilot, training, and validation stages of the study was to provide research equipment and testing costs. Aillis, Inc. was not involved in the clinical study operation, data collection process, auditing, and quality control and assurance of the study. Employees of Aillis, Inc. designed and conducted the study; managed, analysed, and interpreted the data; prepared, reviewed, and approved the article; and were involved in the decision to submit the article.

## Conflict of Interest

SO is the CEO and HK is the CSO of Aillis, Inc. and they hold stock in the company. MF, MS, WT, MIk, and HK are employees of Aillis, Inc. YT and MIw received consultant fees from the company to supervise the study and draft the manuscript.

## Notes

### Funding Statement

This study was funded by the New Energy and Industrial Technology Development Organization (NEDO), Japan (research number 30STS713) and Aillis, Inc..

### Author Declarations

Certified Review Board, Hattori Clinic Clinical Research Ethics Board, Haradoi Hospital, Social Medical Corporation Kobori Central Clinical Research Ethics Committee Medical corporation Takahashi clinic clinical trial examination committee

