## Supplement for "Diagnosing Influenza Infection from Pharyngeal Images using Deep Learning: Machine Learning Approach"

**Supplementary Table 1. List of study sites**

**Pilot stage (37 sites):** Adachi Kyosai Hospital; Ahiko Otolaryngology Clinic; Asahiyama Hospital; Clinic Kashiwanoha; Dozenkai Clinic; Ebisu Clinic; Fukazawa Clinic; Fukuda Clinic (Internal medicine); Funai Ear Nose Throat Clinic; Iguchi Clinic; Irie Child Clinic; Kamei Internal Medicine and Respiratory Clinic; Kamoike ENT Allergy Clinic; Kanagawa Himawari Clinic; Kaneko Clinic; Kanna Hospital; Kawaguchi Kogyo General Hospital; Megumi Clinic; Minami Clinic; Miyazaki RC Clinic; Moriyama Otolaryngology Clinic; Musashino General Hospital; Nishimura Clinic; Nomura Clinic; Okura Otolaryngology Clinic; Rokujizo General Hospital; Ryuto Otolaryngology Clinic; Sawayama Clinic; Shinnakama Hospital; Someya Clinic; Takahashi Clinic; Terada Clinic, Respiratory Medicine and General Practice; Yamagata Clinic; Yamashita Child Clinic; Yasuda Clinic; Yokoyama Children’s Clinic; Yoshimura Child Clinic

**Training stage (64 sites):** Aozora Children’s Hospital; Association of Healthcare Corporation Meiko-kai Ohishi Naika Clinic; Clinic Kashiwanoha; Den-en-tyofu Family Clinic; Dozenkai Clinic; Ebisu Clinic; Eifukuchoekimae Minnano Clinic; Fukuda Clinic (Internal medicine); Himeno Hospital; Ikeda Naika Clinic; Ito ENT Clinic; Kamoike ENT Allergy Clinic; Kanagawa Himawari Clinic; Kikumori Ear, Nose and Throat Clinic; Kimura Clinic; Kumeda Clinic; Kunisaki Makoto Clinic; Maekawa Medical Clinic; Marunouchi Hospital; Mashiki Clinic; Matsuda Pediatric Clinic; Medical Corporation Association Kanwakai Musashikoganei Clinic; Medical Corporation Hitomikai Motomachi Takatsuka Naika Clinic; Medical Corporation Houmankai Umezu Clinic; Medical Corporation Segawa Hospital; Medical Corporation Yuhokai Miho-Clinic; Medical Square Kuhonji Clinic; Megumi Clinic; Miuraiin; Miyanosawa Clinic of Internal Medicine and Cardiology; Miyazaki RC Clinic; Morimoto ENT Clinic; Moriyama Otolaryngology Clinic; Nakamura Cardiovascular Clinic; Nakano Clinic; Nanko Clinic; Nishiyamadou Keiwa Hospital; Nomura Clinic; Okura Otolaryngology Clinic; Primula Clinic; Saino Clinic; Sakata ENT Clinic; Sakura Hospital; Sannou Yamate Clinic; Sasaki Clinic; Sato ENT Clinic; Shimada Clinic; Shirao Clinic of Pediatrics and Pediatric Allergy; Someya Clinic; Sone Clinic Shinjuku; Suzuki Clinic; Suzuki Internal Medicine Clinic; Tanabe Pediatrics; Terada Clinic, Respiratory medicine and General Practice; Ueyama Child Clinic; Umemotokodomo Clinic; Uranishi Clinic; Wada Clinic; Yaesu Clinic; Yamada Clinic; Yamaichi Building Medical Clinic; Yokoyama Children’s Clinic; Yoshimura Child Clinic; YOSHIMURA CLiNiC

**Validation stage (11 sites):** Dozenkai Clinic; Fukuda Clinic (Internal medicine); Himeno Hospital; Kimura Clinic; Nakano Clinic; Sato ENT Clinic; Shimada Clinic; Terada Clinic, Respiratory Medicine and General Practice; Ueyama Child Clinic; Uranishi Clinic; Yaesu Clinic

**Supplementary Figure 1. Feature importance of pharyngeal images and clinical information in the LightGBM model**

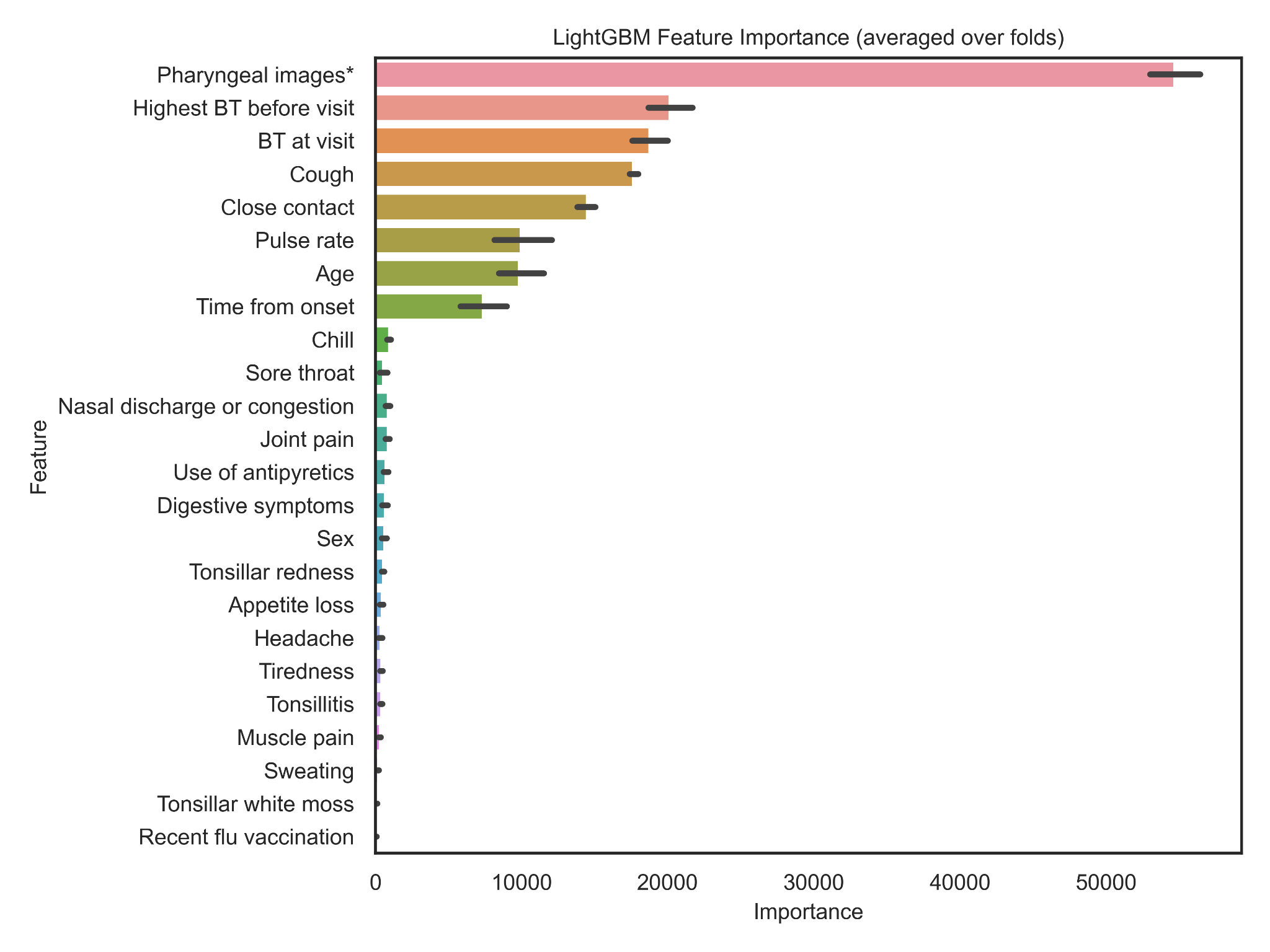

Abbreviation: BT: body temperature

*multi-view convolutional neural network (MV-CNN) influenza probability based on pharyngeal images

**Supplementary Figure 2. Feature importance of pharyngeal images and clinical information in the CatBoost model**

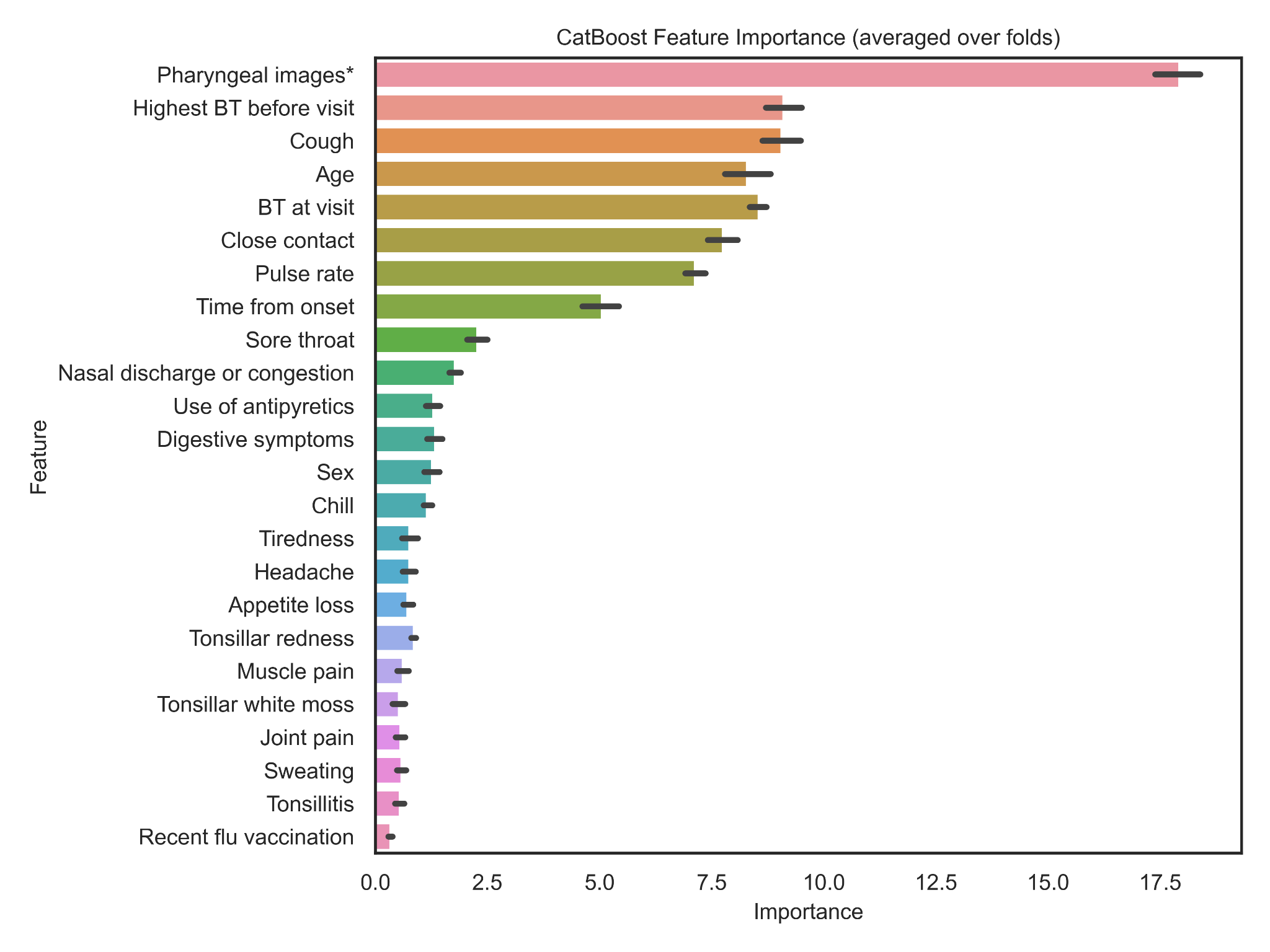
Abbreviation: BT: body temperature

*multi-view convolutional neural network (MV-CNN) influenza probability based on pharyngeal images

**Supplementary Figure 3. Receiver operating characteristic curve of the diagnostic prediction model in the artificial intelligence-assisted diagnosis device**

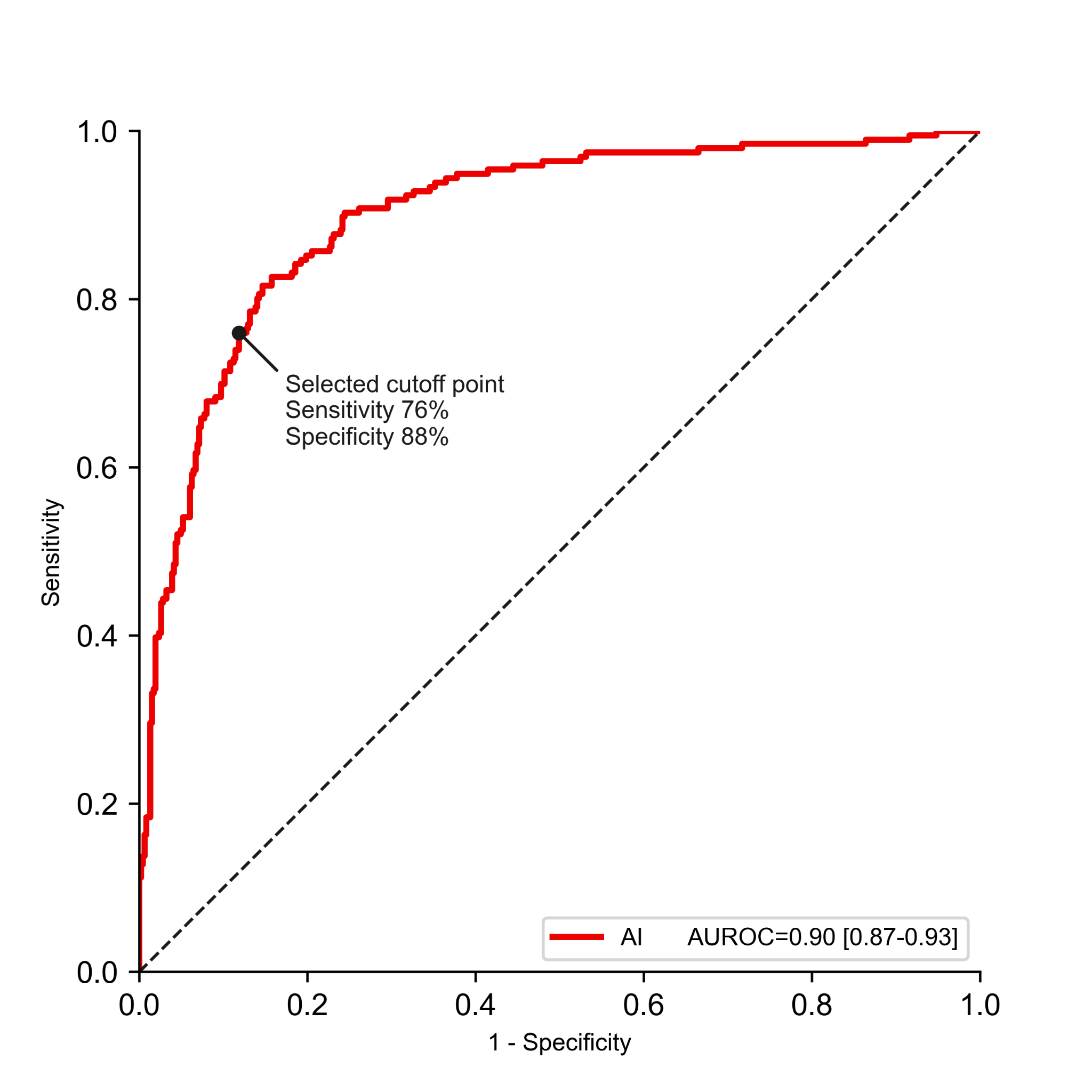

**Supplementary Table 2. Validity of the artificial intelligence-assisted device compared with the gold-standard diagnosis of influenza virus infection based on reverse transcription polymerase chain reaction**

|  | | Influenza virus infection based on RT-PCR | | Total | PPV and NPV  (95% CI), % |
| --- | --- | --- | --- | --- | --- |
|  |  | Positive | Negative |  |  |
| Prediction by the AI-assisted device* | Positive | 149 | 55 | 204 | PPV: 73 (67–79) |
|  | Negative | 47 | 408 | 455 | NPV: 90 (87–92) |
| Total | | 196 | 463 | 659 |  |
| Sensitivity and specificity  (95% CI), % | | Sensitivity:  76 (70–82) | Specificity:  88 (85–91) |  |  |

*According to the selected cut-off point on the receiver operating curve of the diagnostic prediction model of the AI-assisted device shown in Supplementary Fig. 3.

Abbreviations: AI: artificial intelligence, CI: confidence interval, PPV: positive predictive value, NPV: negative predictive value, RT-PCR: reverse transcription polymerase chain reaction
